# Heart-centered positioning and tailored beam-shaping filtration for reduced radiation dose in coronary artery calcium imaging: a MESA study

**DOI:** 10.1101/2021.07.18.21259666

**Authors:** Brendan Colvert, Marzia Rigoli, Amanda Craine, Michael Criqui, Francisco Contijoch

**Author notes:** **Address for Correspondence:** Francisco Contijoch, PhD, 9500 Gilman Drive #0412, La Jolla CA 92093.

## Abstract

**Purpose:** Cardiac CT has a clear clinical role in the evaluation of coronary artery disease and assessment of coronary artery calcium (CAC) but the use of ionizing radiation limits clinical use. Beam shaping “bow-tie” filters determine the radiation dose and the effective scan field-of-view diameter (SFOV) by delivering higher X-ray fluence to a region centered at the isocenter. A method for positioning the heart near the isocenter could enable reduced SFOV imaging and reduce dose in cardiac scans. However, a predictive approach to center the heart, the extent to which heart centering can reduce the SFOV, and the associated dose reductions have not been assessed. The purpose of this study is to build a heart-centered patient positioning model, to test whether it reduces the SFOV required for accurate CAC scoring, and to quantify the associated reduction in radiation dose.

**Methods:** The location of 38,184 calcium lesions (3,151 studies) in the Multi-Ethnic Study of Atherosclerosis (MESA) were utilized to build a predictive heart-centered positioning model and compare the impact of SFOV on CAC scoring accuracy in heart-centered and conventional body-centered scanning. Then, the positioning model was applied retrospectively to an independent, contemporary cohort of 118 individuals (81 with CAC>0) at our institution to validate the model’s ability to maintain CAC accuracy while reducing the SFOV. In these patients, the reduction in dose associated with a reduced SFOV beam-shaping filter was quantified.

**Results:** Heart centering reduced the SFOV diameter 25.7% relative to body centering while maintaining high CAC scoring accuracy (0.82% risk reclassification rate). In our validation cohort, imaging at this reduced SFOV with heart-centered positioning and tailored beam-shaping filtration led to a 26.9% median dose reduction (25-75th percentile: 21.6 to 29.8%) without any calcium risk reclassification.

**Conclusions:** Heart-centered patient positioning enables a significant radiation dose reduction while maintaining CAC accuracy.

## Introduction

Cardiac CT plays a critical role in the non-invasive assessment of cardiovascular disease. For example, coronary artery calcium (CAC) improves risk prediction beyond traditional risk factors^1,2^ and CT coronary angiography (CTCA) is a powerful diagnostic and prognostic tool for coronary artery disease (CAD).^3^ However, the use of ionizing radiation has raised concern about accumulation of radiation dose in the population.^4^ As a result, scan protocols adhere to the “as-low-as-reasonably-acceptable” (ALARA) principle^5^ where dose is reduced by limiting image quality to the minimum needed for the diagnostic task. Improving the risk-benefit ratio of radiation dose is of paramount importance when imaging healthy individuals in screening exams (such as CAC) and when scanning younger patients.^6^ As a result, guidelines often limit the use of CT. For example, CAC screening is only recommended for individuals at intermediate 10-year risk of CAD if traditional tools leave treatment uncertain.^7^

To maximize the risk-benefit ratio of CT imaging, significant efforts and interest in dose-reduction techniques are ongoing.^8^ To reduce dose, patients are placed at the center of the scanner and asked to raise their arms to reduce their cross-sectional extent.^5,9^ In addition, beam-shaping “bow-tie” filters are designed to equalize X-ray photon counts across detectors. By decreasing X-ray fluence to peripheral, less attenuating regions of the body, these filters reduce dose and equalize image quality throughout the field-of-view (FOV).^10–13^ Incorrect patient positioning is known to increase radiation dose^14^ and, consequently, vendors have sought to improve the process by developing automated, camera-based patient position detection^15,16^ and moving tables.^17^

The conventional design of beam-shaping filters and a body-centered patient positioning strategy does not take into account the fact that clinical interest in cardiac imaging is limited to the heart: an off-center subsection of the larger chest cavity. While the entire patient cross-section must be imaged to avoid limited FOV artifacts,^18,19^ it has been shown that significant dose reductions can be achieved by limiting higher X-ray fluence to a smaller subregion.^20–24^ However, the region of interest needs to be positioned at the scanner isocenter and an appropriate beam-shaping filter must be selected. This necessitates determination of the center and diameter of the cardiac region of interest prior to scanning.

In this study, we used the large and diverse MESA study population to define the location and diameter of the heart region based on the distribution of CAC lesions. We show that we can predict the center of the heart region in a patient-specific fashion, using demographic information. By doing so, we can reduce the diameter of the scan field-of-view (SFOV) required for accurate CAC scoring. We applied this approach to contemporary acquisitions at our institution to validate the model predictions of heart positioning and report potential reductions in radiation dose measured via simulation.

## Materials and Methods

### MESA Calcium Lesion Location and Model Prediction

The Multi-Ethnic Study of Atherosclerosis (MESA) studied subclinical atherosclerosis in a diverse cohort of individuals without a history of clinically-recognized cardiovascular disease and its study design has been published previously.^25,26^ The study was approved by the institutional review committee at each participating institution and all subjects gave informed consent.

As part of an approved substudy, calcium scans of 3,151 MESA participants with clinical CAC > 0 on Exam 1 were previously annotated to evaluate the effect of calcium lesion density.^27^ 38,184 calcium lesions were annotated with spatial information (slice number and spatial position in the axial plane), anatomical information (corresponding coronary vessel), along with lesion density (HU) and area (cm^2^). Two calcium scans and their respective clinical scores were also provided. Relevant demographic information for this MESA cohort is shown in Table 1.

**Table 1:**
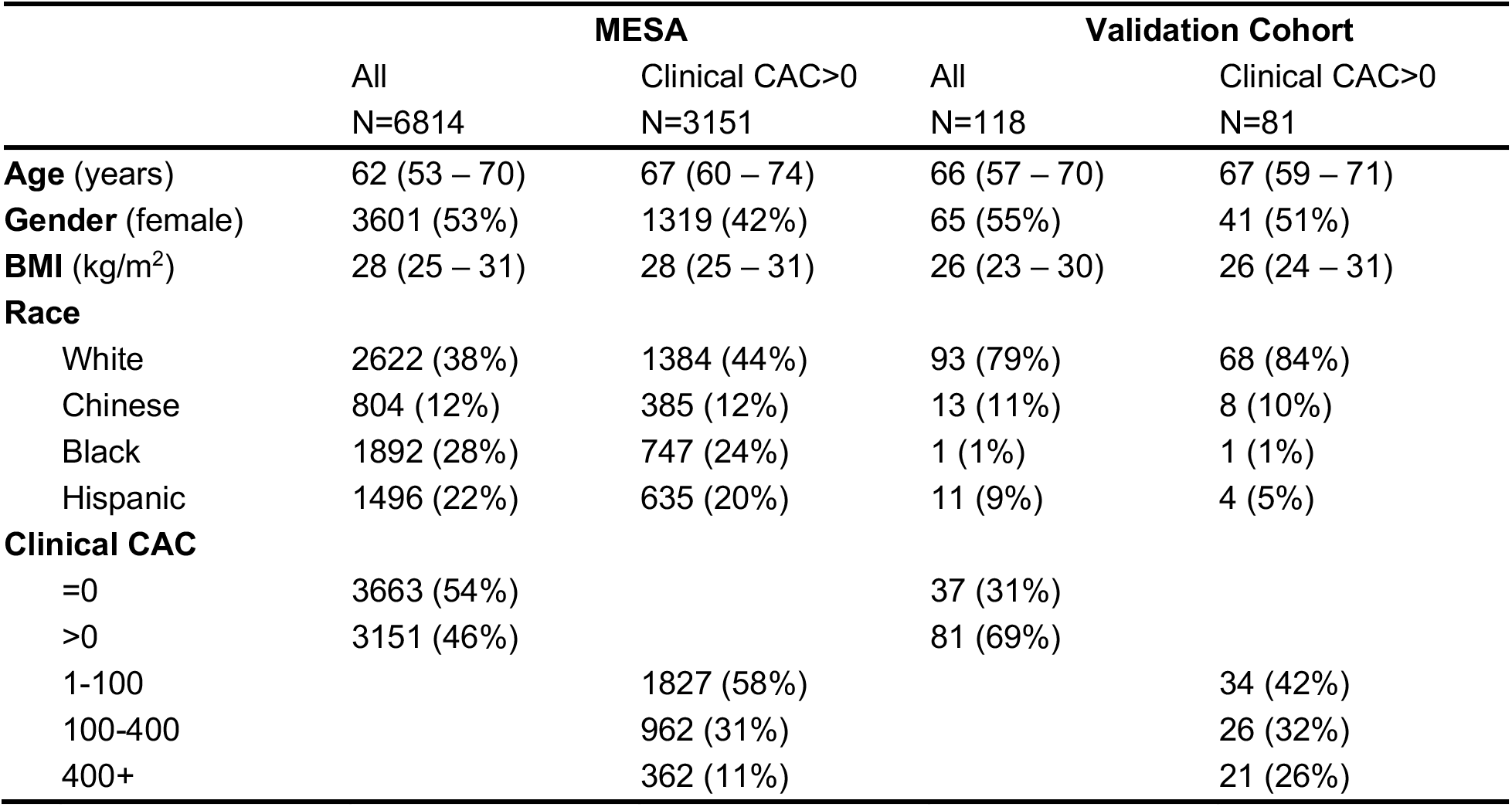
Cohort Demographics. . Relative to the MESA cohort, our validation cohort was similar in age and gender but had significantly lower BMI, included more White participants, and had more coronary artery calcium. Continuous variables are reported as median with the 1st to 3rd quartile range. Categorical values are shown as counts with percentages in parentheses.

To standardize lesion location, a common anatomical origin was defined for each participant. First, images were semantically segmented, as shown in Figure 1, using a deep learning-enabled neural network (as described in Supplemental Methods). From these segmentations, the participant’s midline was defined via the centroid of the spine label. The posterior and anterior skin boundaries at the midline were identified, and the posterior skin extent was defined as the common anatomical origin (Figure 2a). This location was chosen because it is likely to be readily identifiable on scout imaging. The location of each calcium lesion projected to the axial (*x* − *y*) plane was measured relative to this origin. The axial (*z*) lesion coordinates were ignored since our aim was to define a cylindrical SFOV. For each of the four vessels (left main, left anterior descending, left circumflex, and right coronary), a distribution of lesion positions was generated.

**Figure 1:**
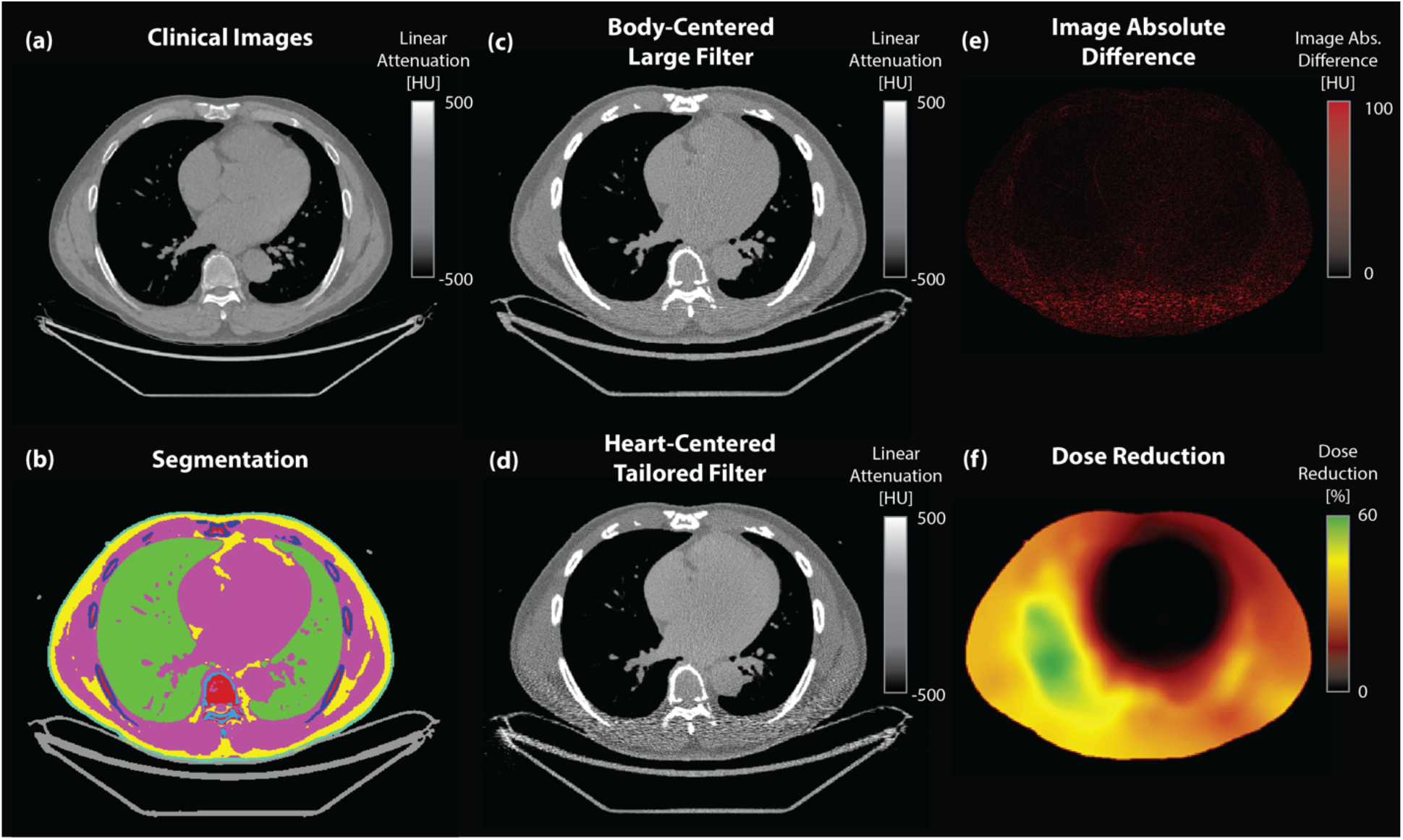
Analysis pipeline. (a) CT images from coronary artery calcium (CAC) scans were analyzed and are depicted here in the central slice (window level (WL)/width (WW) = 0/1000 HU) (b) Automated semantic segmentation via neural network identified tissue types and enabled construction of patient-specific voxelized phantoms (teal: skin, pink: muscle/blood, yellow: fat, light blue: spine, dark blue: bone, green: lung, red: bone marrow, gray: couch). (c) Reconstructed images (WL/WW = 0/1000 HU) were generated via simulation for both (c) the conventional (body-centered, Large filter) and (d) the proposed (heart-centered, tailored filter) approaches. Comparing (e) the absolute difference image (WL/WW = 50/50 HU) and (f) the simulated dose reduction map (WL/WW = 30/30%) illustrates that high image quality is achieved using the tailored approach with significant dose reductions particularly in the lateral and posterior portions.

**Figure 2:**
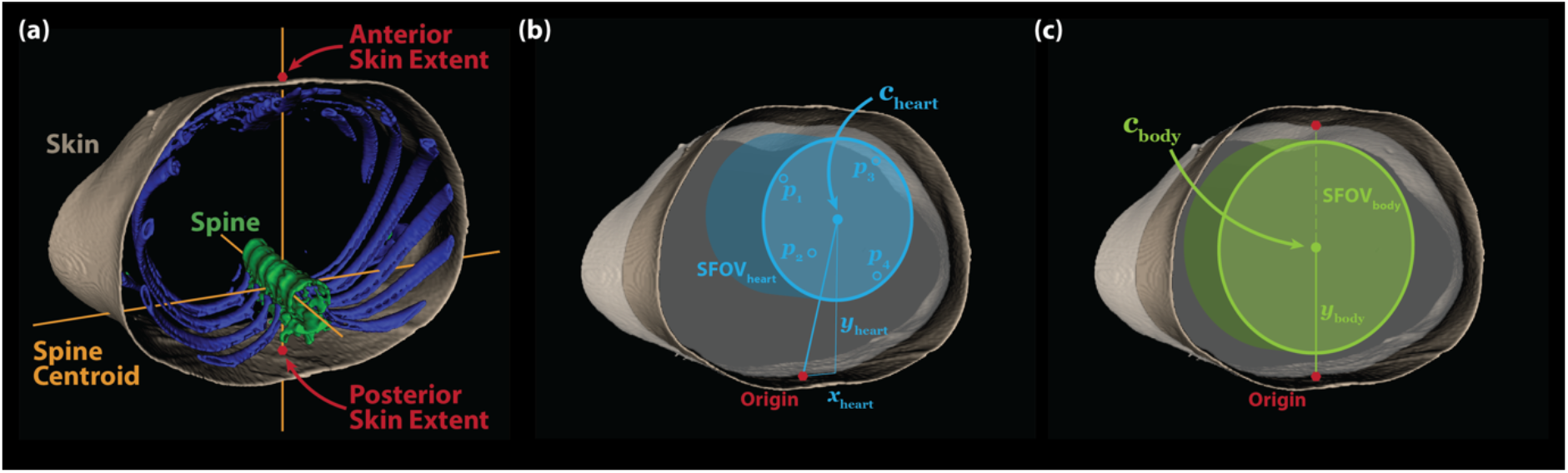
Anatomical Coordinate System and Fields-of-View. (a) The segmented CT images are analyzed to define the patient midline defined by the anteroposterior line passing through the centroid of the spine label. The anterior and posterior skin extent are defined by the intersection of the midline with the skin. (b) The heart center (***c****_heart_*) is determined based on patient-specific, demographic-based prediction of coronary calcium lesion position and is used to define the location of the cylindrical heart FOV. (b) The body center (***c****_body_*) is defined as the midpoint between the posterior and anterior skin extents and is used to define the location of the cylindrical body FOV. The diameters of the scan fields-of-view (SFOV) are determined by CAC accuracy analysis in heart- and body-centered scanning, respectively.

For each of the coronary vessels, a linear regression model was built with demographic inputs (age, sex, BMI, and race/ethnicity) to predict the vessel position in the axial plane. Each regression model (one for each of the four vessels and two axial coordinates, eight linear models in total) was trained by minimizing the mean-square distance between the predicted lesion position from those identified in the MESA dataset. For each patient, the predicted projected vessel positions were denoted ***p****_v_* = (*x_v_*, *y_v_*, 0), where *v* is an index (1 to 4). Heart-centered patient position in the axial plane was found by computing the center of the smallest circle (Figure 2b, cyan dots) that bounds the predicted axial positions of the four coronary arteries.^28^ The heart center position relative to the anatomical origin was denoted ***c****_heart_* = (*x_heart_*, *y_heart_*, 0) and is shown in Figure 2b. The diameter of the SFOV was determined by CAC accuracy analysis with heart centering, as detailed in the next section.

For comparison, a body-centered patient positioning approach was evaluated. The center position was defined as the midpoint between the anterior and posterior skin boundaries at the midline. This position was denoted ***c****_body_* = −0, *y_body_*, 0. and is shown as the green dot on Figure 2c. The diameter of the SFOV was determined by CAC accuracy analysis with body centering, as detailed in the next section.

### Assessment of Calcium Scoring Accuracy with Heart-Centric Positioning and Reduced Scan Field-of-View

To assess the effect of heart-centered positioning compared with body-centering, lesion positions were recomputed relative to ***c****_heart_* and ***c****_body_*, respectively. As the diameter of the cylindrical SFOV was varied, the accuracy of predicted CAC score relative to clinical CAC was assessed. Lesions were only counted towards the predicted CAC score if they were located inside of the SFOV and lesions located outside of the SFOV were not counted. Accuracy was quantified with three metrics: risk reclassification rate (RRR), Lin’s correspondence correlation coefficient (CCC)^29,30^ of log(Agatston+1), and the lesion miss rate (LMR). Risk classification was based on binning predicted CAC scores into the following categories: CAC = 0, 1-100, 100-400, and 400+.^31^ The final values for SFOV_heart_ and SFOV_body_ were chosen such that RRR was approximately 10 times lower than the interscan risk reclassification rate, see Results.

### Validation of Tailored Calcium Scanning in Contemporary Acquisitions

A validation cohort of 156 consecutive calcium scans acquired at our institution between April 2019 and March 2020 on a single CT scanner (Revolution, GE Healthcare, Chicago, IL) with corresponding demographic information were identified with IRB-approved waiver of informed consent to validate the MESA-derived heart-centered positioning model and quantify dose reduction. Images were acquired using the Body filter with ECG gating. 118 individuals met the inclusion criteria of having 1) a full (500 mm diameter) image reconstruction with the entire chest cross-section inside the image (for accurate dose estimation), 2) no implanted metal devices which precluded semantic segmentation and dose estimation, and 3) sufficient image quality for calcium scoring. Relevant demographic information is shown in Table 1. Notably, 81 (69%) individuals had Agatston CAC score > 0.

A certified CTCA imaging cardiologist annotated the calcium scans to generate calcium lesion location data in the same manner as in the MESA substudy. As with the MESA data, individuals were first aligned using the common origin (described above) to account for variation in positioning by the technician. Then, each individual was repositioned in the *x* − *y* plane according to either heart- or body-centered positioning. Heart-centering was evaluated both in terms of calcium scoring accuracy, using the metrics described above, as well as image quality and radiation dose, as described below, via simulation.

### Beam-Shaping Filters

Simulation was used to evaluate changes in radiation dose and image quality due to heart-centered positioning and the use of different beam-shaping filters. The previously described heart-centered positioning approach was combined with three different beam-shaping filters. For beam-shaping filters, two conventional filter profiles were modeled after existing “bow-tie” filters on the GE Revolution scanner,^32^ hereafter referred to as “Large” and “Small”. Additionally, a custom beam-shaping filter profile was designed to deliver the highest X-ray fluence to a 168 mm diameter circular region centered at the isocenter (as determined by the CAC accuracy analysis for heart-centered positioning, see Results). Figure 3 shows the attenuation profiles of the filters under consideration. The cardiac-tailored filter is based on our prior work describing the benefits of more aggressive filtering.^23,24^ The filter profile is designed by convolving a step function at 168 mm between attenuation factors at 80 keV of 0.05 and 1.00 with a Gaussian function (full width at half max ∼100 mm), to avoid imaging artifacts associated with sharp filter edges. The performance of each filter with heart-centered positioning was compared with body-centered positioning and the “Large” filter.

**Figure 3:**
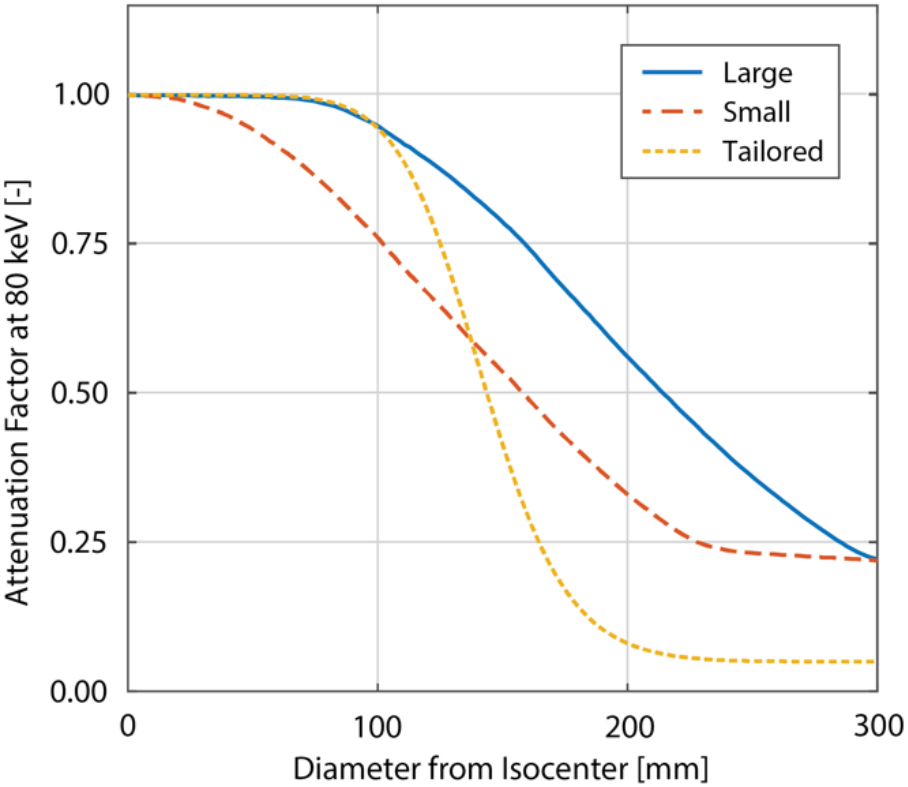
Beam shaping filter profiles. Attenuation profile for filters available on the GE Revolution scanner (Large: solid blue and Small: dashed red) as well as the proposed filter (dotted yellow) with more tailored attenuation profile based on the heart-centered diameter (168 mm) identified in the MESA cohort.

### Radiation Dose and Image Quality Comparison

The absorption of X-ray radiation was simulated volumetrically using voxelized representations of patient anatomies obtained from the semantic segmentations described above. After segmenting the image volume, each tissue label was assigned energy-dependent attenuation *μ*(*x*, *E*) and absorption *μ*_en_(*x*, *E*) coefficients from the NIST table of X-ray Mass Attenuation Coefficients^33^ and commonly accepted values for mass density *ρ*(*x*), where *E* denotes the photon energy and *x* is the spatial coordinate. Following the approach of Bartolac et al.,^34^ photon transport was modeled using the Beer-Lambert Law of Attenuation with a 120 kVp spectrum *φ_o_*(*E*) and dose was approximated with collision kernma. This calculation was implemented in MATLAB (R2019b, Mathworks, Natick, MA), for more details on the simulation, see Supplemental Methods. Since absorbed dose is proportional to the tube current *I*, we quantified the dose rate denoted *U*(*x*) with units of mGy · mA^-1^. A dose simulation was performed for each patient (n=118), with four filter and position combinations, leading to a total of 472 simulations. Mass density-weighted spatial dose averages were computed in four anatomical regions of interest: the entire thorax (*U*_thorax_), the lungs (*U*_lung_), the spine (*U*_lung_), and the skin (*U*_skin_).

In addition to changing radiation dose, repositioning changes the X-ray fluence through the heart region of interest and therefore the noise in the reconstructed image. The imaging process was simulated at the central slice of the patient-specific voxelized phantoms. Poisson noise was incorporated at nine fluence levels with three independent realizations to generate 27 sinograms with various noise levels as well as one noiseless ground truth sinogram. This led to 28 sinograms x 118 patients x 4 conditions with a total of 13,216 realizations. For details of the simulation, see Supplemental Methods. Each sinogram was reconstructed using fanbeam filtered backprojection^35^ with a Ram-Lak filter and the error image was computed as the difference between the noisy realization and the noiseless reconstruction. Noise was quantified according to both the mean square error **e*_MSE_* as well as the 95^th^ percentile of the squared image error *e*_95_. Owing to the Poisson law, it is expected that *e*^2^ ∝ *I*^-1^ thus, for each patient, filter, and positioning combination, the relationship between image noise and tube current was fit using linear regression. The slope *m* of the fitted curve has units of mA · HU^2^ and quantifies the effects of scanning mode on image quality.

For the case of constant fluence, we computed reductions in dose (ratios of *U*_thorax_ between scan modes), and reductions in image noise (ratios of *m* between scan modes). Since patient positioning and the beam-shaping filter impact both image noise and dose simultaneously, we computed a figure of merit *FOM* = *Um* which quantifies the constant-noise dose and has units of mGy · HU^2^. This enables a fair comparison by computing ratios of *FOM* between scan modes and is a true measure of dose reduction for equivalent image quality. Regional dose reductions are reported in this constant-noise condition.

### Statistical Analysis

Measures were tested for normality using the Shapiro-Wilks test. Unless otherwise specified, normally distributed variables are reported as mean ± 0.674*standard deviation (to match quartiles of the normal distribution). Measures where the normality hypothesis was rejected are reported as median with first and third quartiles (Q1 and Q3, respectively). Student’s t-test and ANOVA were used to assess statistical significance for normally distributed variables while Wilcoxon rank-sum and Kruskal-Wallis were used to assess statistical significance for non-normally distributed variables, all at significance level p = 0.05.

Rate metrics (RRR and LMR) observed in the validation cohort were assessed using post-hoc power analysis. Confidence bounds for the observed MESA values were determined for 80% power and significance level 0.05, and accounting for the size of the validation cohort. Validation rate metrics observed outside of these bounds permitted rejection of the null hypothesis that the validation data were the same as those from MESA.

Multivariate LASSO linear regression of dose reduction was performed to identify associations between dose reduction and image acquisition parameters (bowtie filter and positioning approach) and demographics.

## Results

### Heart-Centered Patient Positioning from MESA Calcium Lesions

In the MESA population, the body center position was found to be *y*_body_ = 127 mm (Q1-Q3: 117 to 137 mm) anterior relative to the posterior midline location (anatomical origin). Patient-specific demographic modeling of the heart position led to a *y*_heart_ = 159 mm (Q1-Q3: 148 to 169 mm) anterior and *x*_heart_ = 12 mm (Q1-Q3: 9 to 15 mm) leftward shift. Four demographic variables (age, sex, BMI, and race/ethnicity) were utilized, and each was a significant (p<0.05) predictor in at least one vessel/coordinate model.

### Impact of Heart-Centered Positioning on Calcium Scoring in MESA

Heart-centered positioning decreased the SFOV needed for high accuracy CAC scoring (Table 2 and Figure 4). Repeat calcium scoring in MESA had an 8.2% risk reclassification rate (RRR). For a RRR of 0.86% (approximately ten times lower than repeat scanning), heart centering reduced the required SFOV 25.7%. In the MESA cohort, RRR = 0.86% was associated with a very small number of missed lesions (1.91%) and very high concordance correlation coefficient (0.996) for Agatston scoring.

**Table 2.** Heart centering enables assessment within a reduced scan field-of-view with high calcium scoring accuracy as assessed in both the MESA and validation cohorts. RRR: Risk Reclassification Rate. CCC: Concordance Correlation Coefficient for log(Agatston+1) scoring. LMR: Lesion Miss Rate. Confidence bounds for RRR and LMR were computed using post-hoc power analysis based observed MESA values, 80% power and significance level 0.05, and accounting for the size of the validation cohort (N=81 individuals and N=979 lesions, respectively). Asterisks indicate where validation performance was better than the predicted confidence bound.

|  | Body-Centered | Heart-Centered |
| --- | --- | --- |
| Risk Reclassification Rate (%) |  |  |
| MESA | 0.86 (0.00 – 4.87) | 0.86 (0.00 – 4.87) |
| Validation | 0.00 | 0.00 |
| Concordance Correlation Coefficient |  |  |
| MESA | 0.995 | 0.996 |
| Validation | >0.999 | >0.999 |
| Lesion Miss Rate (%) |  |  |
| MESA | 1.75 (0.71 – 3.04) | 1.91 (0.81 – 3.25) |
| Validation | 0.31* | 0.31* |

**Figure 4:**
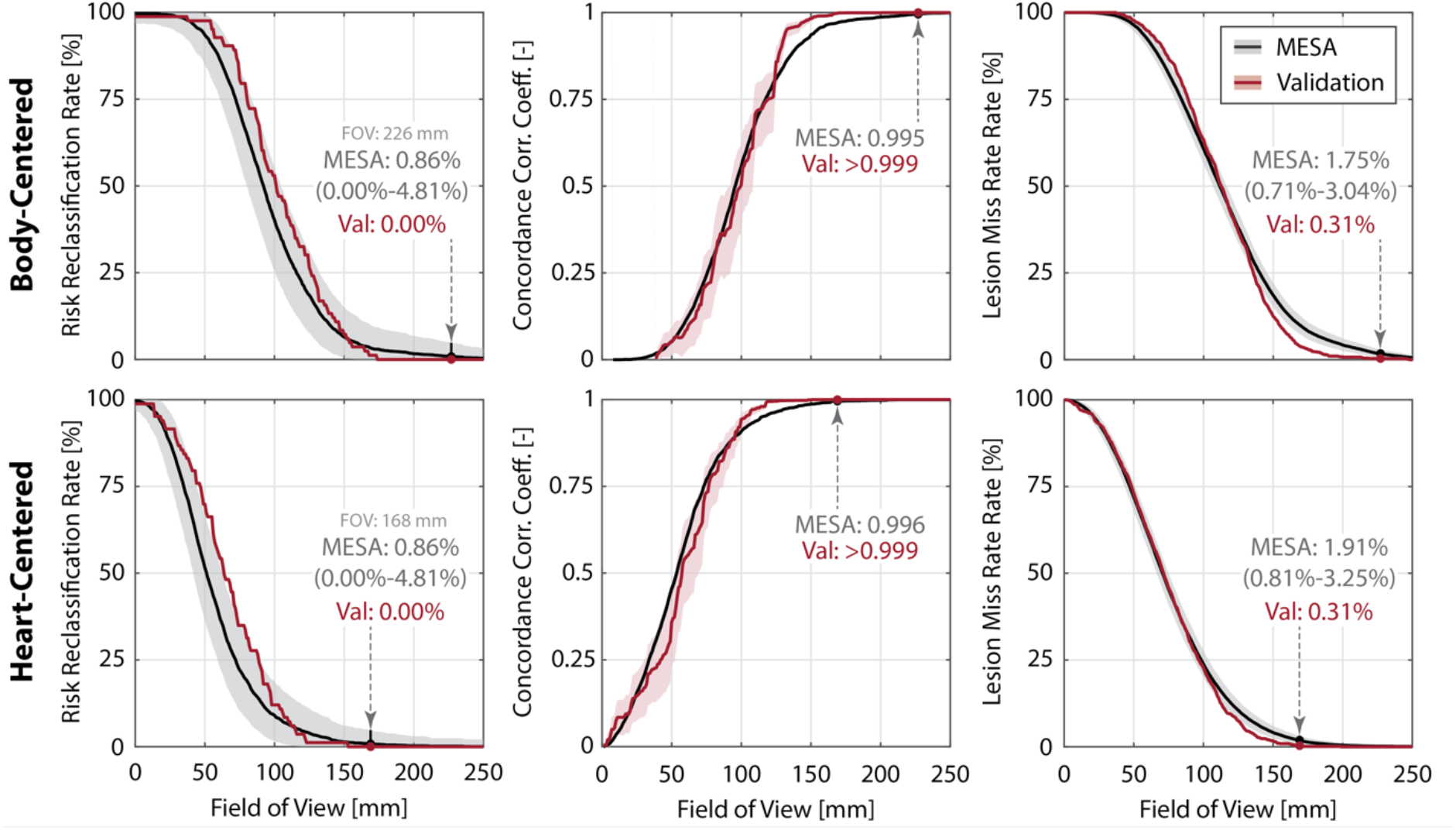
Heart-centering correctly captures calcium lesions and permits accurate Agatston scoring at reduced scan field-of-view (SFOV). Heart centering (top row) reduces the SFOV relative to body-centering (bottom row) while maintaining high calcium score risk classification accuracy (left), high concordance correlation coefficient (middle), and low lesion miss rate (right). MESA data is shown in black with the resulting performance in the validation cohort shown in red. Confidence bounds for RRR and LMR were computed using post-hoc power analysis based observed MESA values, 80% power and significance level 0.05, and accounting for the size of the validation cohort (N=81 individuals and N=979 lesions, respectively).

### Validation of Heart-Centered Positioning and Calcium Scoring

Positioning and calcium scoring accuracy (with MESA-derived SFOV diameters) were validated in 118 individuals who underwent calcium scanning at our institution. As shown in Table 1, the validation cohort was of similar age (All: p=0.07, CAC>0: p=0.28) and gender (All: p=0.63, CAC>0: p=0.11) as the MESA cohort but had significantly lower BMI (All: p<0.01, CAC>0: p=0.05), lower prevalence of Black and Hispanic subjects (All: p<0.01, CAC>0: p<0.01), and higher CAC scores (higher percentage of individuals with CAC>0 as well as scores > 400, p <0.01).

Body centering in our validation cohort was found to be *y*_body_ = 120 mm (Q1-Q3: 110 to 131 mm) anterior relative to the posterior midline location. The heart-centered positioning model predicted a *y*_heart_ = 156 mm (Q1-Q3: 145 to 164 mm) anterior and *x*_heart_ = 12 mm (Q1-Q3: 10 to 14 mm) leftward shift. In both patient positioning approaches, we observed perfect calcium risk classification (RRR = 0.00%), high calcium scoring accuracy (CCC > 0.999), and lesion miss rates below the range predicted by MESA, as shown in Figure 4. This confirms the ability of the heart-centered positioning model to reduce the FOV 25.7% relative to body centering without the loss of calcium scoring accuracy.

### Effect of Heart-Centering and Tailored Bow-Tie Filters on Dose and Image Noise

The distributions of simulation-predicted dose reductions (relative to body-centered imaging with the Large filter) for heart centered positioning with different beam-shaping filters are shown in Figure 5. Summary statistics are shown in Table 3. For both noise reduction (*m*) and constant-image dose reduction (*FOM* = *Um*), we observed no significant differences (p>0.05) between quantifying noise with the mean squared error (*e_MSE_*) and the 95^th^ percentile squared error (*e*_95_), therefore we report all metrics below using *e_MSE_*. Heart centering with tailored filtering reduced *FOM* by 26.9% (Q1-Q3: 21.5 to 29.8%). The improvement is primarily due to the filter reducing *U* (25.5%, Q1-Q3: 20.4 to 29.4%) with a small improvement in *m* (0.6%, Q1-Q3: −1.1 to 2.1%). The use of the Small filter reduced *FOM* 12.1% (Q1-Q3: 7.7 to 16.4%) with heart centering. The reductions in *U* (19.4%, Q1-Q3: 15.8 to 22.0%) are partially offset by an increase in *m* (7.9%, Q1-Q3: 5.0 to 11.0%). All changes in dose or noise were significantly different from zero (p<0.05) except the change in MSE using the Tailored filter (p=0.06). Reductions in dose or image noise were not significantly different between individuals with CAC = 0 and those with CAC > 0 (p>0.05 for all conditions).

**Figure 5:**
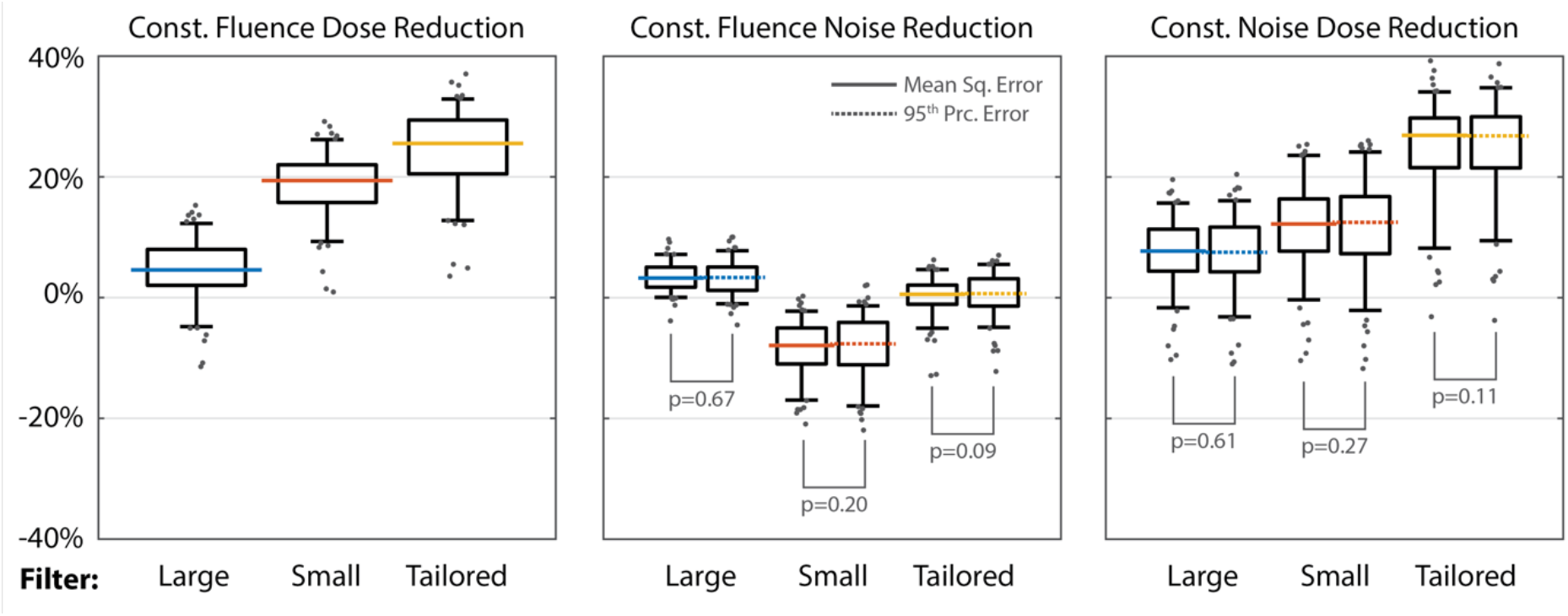
Dose reductions, improved image quality, and combined effects in the validation cohort. The tailored beam-shaping filter (yellow) provides a 25.5% (Q1-Q3: 20.5-29.4%) reduction in constant-fluence dose. The Small filter (red) enables a reduction of 19.4% (Q1-Q3: 15.8-22.0%) in constant-fluence dose, but experiences a 7.9% (Q1-Q3: 5.0-11.0%) increase in constant-fluence noise. Combined, these effects lead to a graded improvement, with the Tailored filter exhibiting a 26.9% reduction in constant-image-noise dose (Q1-Q3: 21.6-29.8%). Horizontal lines indicate median value, boxes indicate first and third quartiles, whiskers indicated 5^th^ and 95^th^ percentiles. Gray circles show outliers. We find no significant differences between mean and 95^th^ percentile squared error-based noise quantification methods (p>0.05).

**Table 3:** Heart-centering enabled tailoring of the acquisition enables a reduction in dose. Mean (or median where appropriate) values shown with ranges from 1^st^ to 3^rd^ quartiles. Simulations predict a 26.9% constant-image-noise dose reduction can be achieved via heart centering with tailored beam-shaping filtration. Use of Small filter decreases constant-noise dose 12.2% when accounting for degraded image noise (7.9% increase) and lowered dose (12.2% decrease). Dose and noise reductions found to be significantly different from zero (p<0.05) are marked with an asterisk. No significant differences (p>0.05) were found between MSE and 95^th^ percentile noise accounting methods.

|  | Large | Small | Tailored |
| --- | --- | --- | --- |
| **Constant Fluence** |  |  |  |
| Dose Reduction (%) | 4.6 (2.0 – 8.0)* | 19.4 (15.8 – 22.0)* | 25.5 (20.5 – 29.4)* |
| MSE Reduction (%) | 3.3 (1.8 – 4.8)* | -7.9 (-11.0 – -5.0)* | 0.6 (-1.1 – 2.1) |
| 95<sup>th</sup> Prc. Err. Reduction (%) | 3.2 (1.4 – 5.1)* | -7.7 (-11.1 – -4.1)* | 0.6 (-1.4 – 3.1)* |
| Noise Method Significance | $p=0.67$ | $p=0.20$ | $p=0.09$ |
| **Constant Image Noise** |  |  |  |
| Dose Reduction, Const. MSE (%) | 7.7 (4.4 – 11.3)* | 12.2 (7.7 – 16.4)* | 26.9 (21.5 – 29.8)* |
| Dose Reduction, Const. 95<sup>th</sup> Prc. Err. (%) | 7.5 (4.3 – 11.7)* | 12.5 (7.3 – 16.8)* | 26.8 (21.5 – 30.0)* |
| Noise Method Significance | $p=0.61$ | $p=0.27$ | $p=0.11$ |

### Predictors of Calcium Score Accuracy and Dose Reduction

The positioning model, Small and Tailored beam-shaping filter, BMI, sex, and Agatston score were significantly associated with dose reduction (overall model adjusted R^2^ = 0.72). Heart centering (β = 7.4, CI: 5.7 to 9.2, p<0.001), small filter (β = 4.2, CI: 2.4 to 6.0, p<0.001), tailored filter (β = 17.1, CI: 15.3 to 18.8, p<0.001), BMI (β = 0.3, CI: 0.2 to 0.4, p< 0.001), and male sex (β = 2.9, CI: 1.6 to 4.1, p<0.001) were associated with improved dose reduction while log(Agatston+1) was associated with decreased dose reduction (β = −0.6, CI: −0.9. to −0.2, p<0.001).

### Regional Dose Differences

Tailored beam filtration and heart centering reduced constant-noise dose 35.7% to the spine (Q1-Q3: 30.4 to 41.2%), 26.4% to the lungs (Q1-Q3: 21.9 to 31.1%), and 9.8% to the skin (Q1-Q3: 1.8 to 14.8%), see Table 4. Use of the Small filter led to reductions in lung and spine dose (16.3 and 31.1%, respectively) but an increase in skin dose (17.2%, Q1-Q3: 12.4 to 22.9%).

**Table 4:**
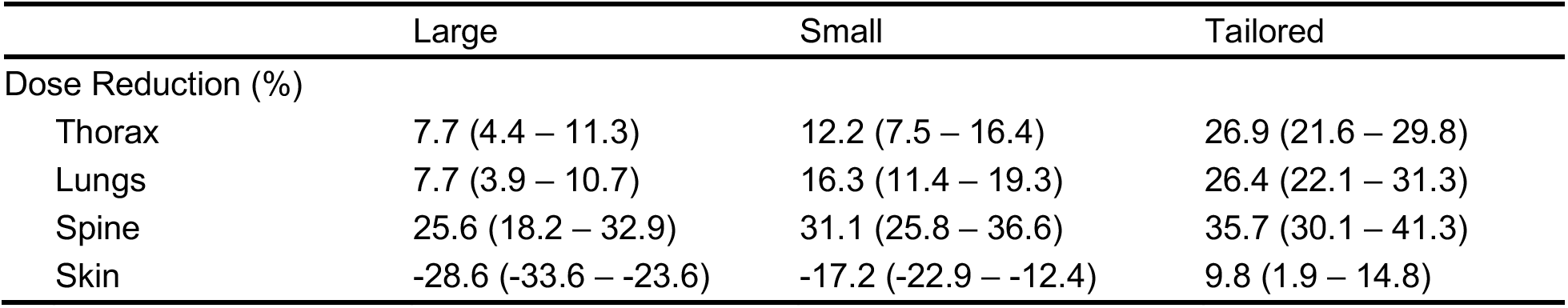
Constant-image-noise dose reduction shows regional variations. Median values shown with ranges from 1^st^ to 3^rd^ quartiles. Heart centering with tailored beam-shaping filtration decreases simulation-predicted dose to all four anatomical regions. The Small filter reduces dose to the lungs and spine but leads to an increased skin dose.

|  | Large | Small | Tailored |
| --- | --- | --- | --- |
| Dose Reduction (%) |  |  |  |
| Thorax | 7.7 (4.4 – 11.3) | 12.2 (7.5 – 16.4) | 26.9 (21.6 – 29.8) |
| Lungs | 7.7 (3.9 – 10.7) | 16.3 (11.4 – 19.3) | 26.4 (22.1 – 31.3) |
| Spine | 25.6 (18.2 – 32.9) | 31.1 (25.8 – 36.6) | 35.7 (30.1 – 41.3) |
| Skin | -28.6 (-33.6 – -23.6) | -17.2 (-22.9 – -12.4) | 9.8 (1.9 – 14.8) |

## Discussion

Our study used the large and diverse MESA study population to define the location and diameter of the heart region based on the distribution of CAC lesions. Heart-centered positioning enabled a 25.7% reduction of the SFOV without loss of calcium scoring accuracy. We validated the heart-centered positioning model and predicted that moderate (12.1%) reductions in constant-image-noise dose are possible with the Small beam-shaping filter and that a significantly higher reduction (26.9%) could be obtained with the use of a tailored filter with more aggressive peripheral beam attenuation.

Our lesion prediction model is built using the large and demographically diverse MESA cohort. We expect this will improve its ability to generalize to a broader set of patient populations. Our validation study in a contemporary clinical cohort provides evidence for the utility of the model in a modern clinical population. Our study is further strengthened by the use of automated tools to identify anatomical features of interest for positioning and quantify dose. Further, we used a polyenergetic, volumetric CT simulation, to ensure that our predicted dose reductions are as accurate as possible. The tools used in this manuscript, including the automated segmentation which enables identification of the common anatomical origin as well as the predictive model for the heart center, are available upon reasonable request to the corresponding author.

We predict dose reductions and imaging accuracy in CAC imaging with currently available filters, but several other CT imaging protocols could also benefit from our heart centering approach. The region we identified is likely very close to the region of interest in coronary angiography. However, our region of interest was defined based on the presence of calcified lesions. Therefore, future studies aim to assess whether our model captures the presence of non-calcified stenosis and disease in distal artery vessels. Additionally, other cardiac applications such as the evaluation for transcatheter aortic valve replacement could benefit from a tailored acquisition with slight modification of the region of interest. In addition, the congenital and pediatric cardiac population often undergo repeated CT imaging for diagnosis and follow-up and would benefit greatly from dose reduction. More generally, we anticipate that non-cardiac applications could benefit from this general approach if a reliable method is available to identify and define the region of imaging interest.

While we show that heart-centering enables a reduced SFOV, current X-ray beam-shaping filters are not designed for imaging of a smaller region of interest. While currently available filters could be used to realize dose reductions (12.2% with the Small on the GE scanner), we calculate that a more tailored approach could enable larger reductions (26.9% for our proposed profile). The tailored filter profile we propose is conservative relative to prior designs suggesting further reductions could be realized. Finally, given the design of current commercially-available filters, we evaluated a tailored filter with a single effective diameter. Additional reductions can be achieved if the SFOV were adjusted for each individual, but this would require a variable-diameter filtering approach. Such designs are the subject of ongoing research^21,36^ and may enable further tailoring of the X-ray beam and associated reductions in dose.

Our findings represent a conservative estimate of how much the SFOV can be reduced. Specifically, in our assessment, we assumed all lesions outside of the desired SFOV would not be detected. However, as shown in Figure 1, while there is additional noise outside the prescribed SFOV, the image may still be of sufficient diagnostic quality for evaluation. Additionally, the shape of the tailored filter is smooth allowing for some imprecision in patient positioning. Therefore, additional dose reductions (either by more aggressive reduction of peripheral fluence or a smaller SFOV) can likely be realized in practice. However, lesion detectability is a task which is beyond the scope of this study. Further, we defined our SFOV targeting a risk reclassification rate an order of magnitude lower than the observed MESA interscan reproducibility to minimize bias from our approach. We did not observe any misclassifications in our validation cohort which implies more aggressive reductions in the SFOV may be possible while remaining below the scan reproducibility threshold.

In some applications, cardiac scans are performed longitudinally. Prior scans could be used to further tailor the imaging approach we describe. For example, imaging of a specific coronary vessel (to assess stent patency, for example), could be performed with a SFOV even more limited than the one used to assess the entire heart. However, realizing additional reductions would require a filter that could adapt to the desired SFOV.

Our approach to patient positioning was developed with implementation on contemporary scanning equipment and the constraints of our data in mind. The design goal was to predict *where* calcium lesions are relative to anatomical landmarks which could be reliably identified and localized with respect to the scanner coordinates. Specifically, we chose the posterior skin extent as an anteroposterior landmark because it corresponds to where the patient meets the table surface. We chose the spine as a lateral landmark as it can be readily identified on scout imaging, or in some cases externally. These landmarks were also readily identified by our neural network segmentation of the calcium images in our datasets. We anticipate that more patient-specific predictions of lesion position could be made directly from scout imaging, the implementation of which is the subject of future study in our lab.

Our findings have several limitations. First, while we demonstrated significant dose reductions could be achieved with a tailored filter profile, our results are based on simulation. We modeled our approach after the work of Bartolac et al.^34^ who used the same method to estimate dose when tailoring the X-ray beam to a region of interest. Approximating the dose by collision kerma accounts for the volumetric, polyenergetic absorption of X-ray radiation, but omits scatter. In the diagnostic X-ray energy range, scatter accounts for a significant fraction of total absorbed dose.^37–39^ This effect will impact estimates of spatial dose distribution both in the baseline approach as well as with the tailored filter. It is important to acknowledge that a nontrivial amount of scatter from the high fluence region at the center of the filter may lead to dose deposited outside of the desired FOV. A common approach to incorporate scatter in dose models is a convolution-superposition model,^40^ where energy from the primary interaction is spread over a larger spatial area. The energy deposition kernel which describes this phenomenon suggests that the amount of energy attributable to scatter is isotropic in the diagnostic X-ray energy range and falls of rapidly away from the primary interaction site.^41,42^ We believe that our method provides reasonable first-order estimates of ratios of dose between scanning modes and that our predicted trends will hold when we validate our results using more resource-intensive Monte Carlo simulations that incorporate scatter in future studies.

Second, although some newer scanners allow lateral table movement, most scanners in clinical use do not have this capability. For scanners without lateral table movement, the offset could be applied when the patient lays on the table since the midline could be estimated externally. Furthermore, we found that the required lateral shift from the midline for heart-centered scanning was small (median 12mm leftward) and varied little between patients (Q1-Q3: 9 to 15 mm). For this reason, a precise lateral shift is less important for heart-centered scanning as compared to the required anteroposterior shift which is much larger (median 159 mm) and more variable between patients (Q1-Q3: 148 to 169 mm). Thus, our study suggests that while lateral table movement would improve positioning, anteroposterior table movement is more critical and is widely available on current scanners.

Third, we acknowledge that there are practical design and manufacturing constraints which limit the ability to deploy a new beam-shaping filter in clinical scanning equipment. However, cardiac imaging is an important and growing application of CT to the point that some scanners are being designed specifically around this clinical task,^43^ so we believe that it is valuable to assess cardiac-specific filter designs.

Lastly, while we validated the demographic-based patient positioning model in individuals scanned at our institution almost 20 years after the MESA images were acquired, our algorithm was applied retrospectively. A prospective study validating the accuracy and dose reductions is planned as future work.

## Conclusions

In this study, we used the large and diverse MESA study population to define the location and diameter of the heart region based on the distribution of CAC lesions. We built a demographic-based model for patient-specific heart position prediction. Then, we demonstrated that the accuracy of CAC scoring can be maintained while centering the heart at the scanner isocenter and reducing the scan field of view. We validated our findings in a contemporary clinical cohort and predicted that significant reductions in radiation dose could be achieved when heart-centered scanning was paired with a tailored, aggressive beam-shaping filter. The results of this study show that heart-centered patient positioning has the potential to reduce the required scan field-of-view for calcium scoring as well as other cardiac CT imaging applications, enabling dose reduction via tailored beam shaping filtration.

## Data Availability

Data and tools are available upon reasonable request to corresponding author.

## Acknowledgments

The MESA study was supported by contracts 75N92020D00001, HHSN268201500003I, N01-HC-95159, 75N92020D00005, N01-HC-95160, 75N92020D00002, N01-HC-95161, 75N92020D00003, N01-HC-95162, 75N92020D00006, N01-HC-95163, 75N92020D00004, N01-HC-95164, 75N92020D00007, N01-HC-95165, N01-HC-95166, N01-HC-95167, N01-HC-95168 and N01-HC-95169 from the National Heart, Lung, and Blood Institute, and by grants UL1-TR-000040, UL1-TR-001079, and UL1-TR-001420 from the National Center for Advancing Translational Sciences (NCATS). The authors thank the other investigators, the staff, and the participants of the MESA study for their valuable contributions. A full list of participating MESA investigators and institutions can be found at http://www.mesa-nhlbi.org.

Analysis of the calcium lesion locations was supported by NIH-NHLBI R01 HL116395. BC received other research support from the NVIDIA Corporation. FC is supported by NIH NHLBI K01HL143113. The authors would like to acknowledge Dr. Elliot R. McVeigh for advice and guidance during the study and BC thanks ERM for financial support.

## Supplemental Methods

### Supplemental Methods 1: Neural Network Segmentation

A deep learning approach was used to accomplish semantic segmentation of CT images. For training purposes, 9 slices equally spaced in 1 cm increments (the center slice and 8 in the superior-inferior direction) from noncontrast chest CT image volumes in 30 individuals, 15 female and 15 male, reconstructed at 2.5 mm slice resolution (total of 270 slices in the training dataset) were manually segmented using ITK-SNAP.^1^

A conventional neural network approach for medical image segmentation (U-Net)^2^ was utilized where the encoder-decoder architecture uses a series of repeated convolutions with downsampling and upsampling to learn salient features in the image which lead to accurate semantic segmentation. We used a U-net with four downsampling and upsampling layers and powers of two for feature depth in each convolutional layer, as depicted in Figure S1. Our U-Net was developed and trained in PyTorch.

The neural network was evaluated using a cross-entropy loss function and was optimized using RMSProp. Five-fold cross validation was performed by splitting the training dataset into 5 subgroups, each with 3 females and males. The networks were optimized using a batch size of 8 slices and were trained for 80 epochs. Mean training and validation loss were logged over the course of training at the end of each epoch and the state of the network was saved to disk. From the five networks and 80 epochs, the network that had the lowest validation loss over the course of training was selected as the model to be used for segmentation in further analysis.

### Supplemental Methods 2: Patient-Specific Dose and Imaging Simulation

Let µ(*x*, *E*) be the linear attenuation field where *x* is the spatial coordinate and *E* is the photon energy. This linear attenuation field is obtained by semantic segmentation of clinical chest CT images (see Figure S2). Each tissue label was assigned linear attenuation and absorption coefficients derived from the NIST table of X-ray attenuation coefficients^3^ and commonly-accepted values for mass density. Also, let ψ(γ) be the beam shaping filter attenuation factor where γ(*x*) is the angle in the axial plane between *x* and the central ray on the detector and let *d*(*x*, β) be the distance from the X-ray source where β is the gantry angle, as shown in Figure S2. Then, the photon flux rate is

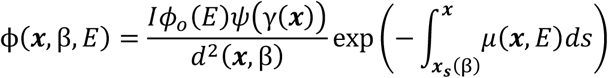

where I is the tube current, *φ_o_*(*E*) is the source spectrum (in photon counts per mA), and *x_s_*(*β*) is the source position. The exponential term is the ‘projection’ as shown in Figure S2.

Let *μ*_en_(*x*, *E*) and *ρ*(*x*) be the linear absorption and mass densities, respectively. The rate of dose absorption (approximated by collison kerma, neglecting scatter) is then given by

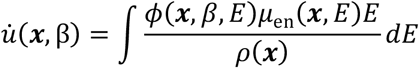

This quantity is depicted as the ‘view dose’ in Figure S2. Finally, to compute the dose over a complete gantry cycle, we integrate view dose for a full set of views

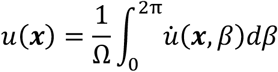

where Ω is the gantry rotation rate. These integrals are accomplished via numerical integration. Where appropriate, dose is spatially averaged with density weighting. Let ℛ denote a region in space, then

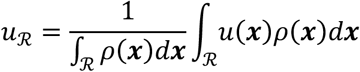

is the regional dose average.

Since radiation dose *u* is linearly proportional to X-ray tube current *I*, dose was quantified by computing the amount of absorbed dose per mA, denoted *U* = *u*⁄*I*.

A similar procedure was performed to simulate the imaging process. This process is depicted in Figure S3. First, the linear attenuation field is forward projected onto a fan-beam detector geometry

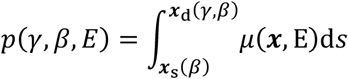

where *x*_d_(*γ*, *β*) is the detector position. The expected number of photons detected at each detector position is given by

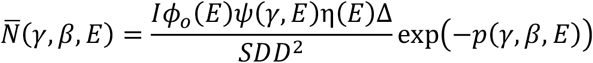

where *SDD* is the source-detector distance, *η*(*E*) is the detector efficiency, and Δ is the detector area. The true (stochastic) number of detected photons is determined by a realization of a Poisson random variable *N* = Poisson(*N*^L^). Similarly, the air scan (no patient in the scanner) is given by

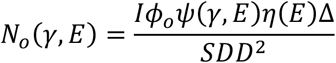

The sinogram can then be computed as

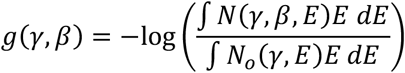

Images are then reconstructed using fanbeam filtered backprojection with a Ram-Lak filter.

Noise levels are varied by selecting different values for the tube current. We computed the difference between the noisy realization and the noiseless reconstruction to measure the error. Then, we quantified the noise using both the mean squared error 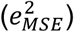 and the 95^th^ percentile squared error 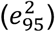 within the region of interest. Owing to the Poisson law, we expect that *e*^2^ *−I*^-1^. This is illustrated in Figure S4. In our analysis, we use linear regression to fit a line to the relationship between tube current and image noise. The slope of the fit *m* is used to quantify the effect of scanning mode on image quality.

## Supplemental Figures

**Figure S1:**
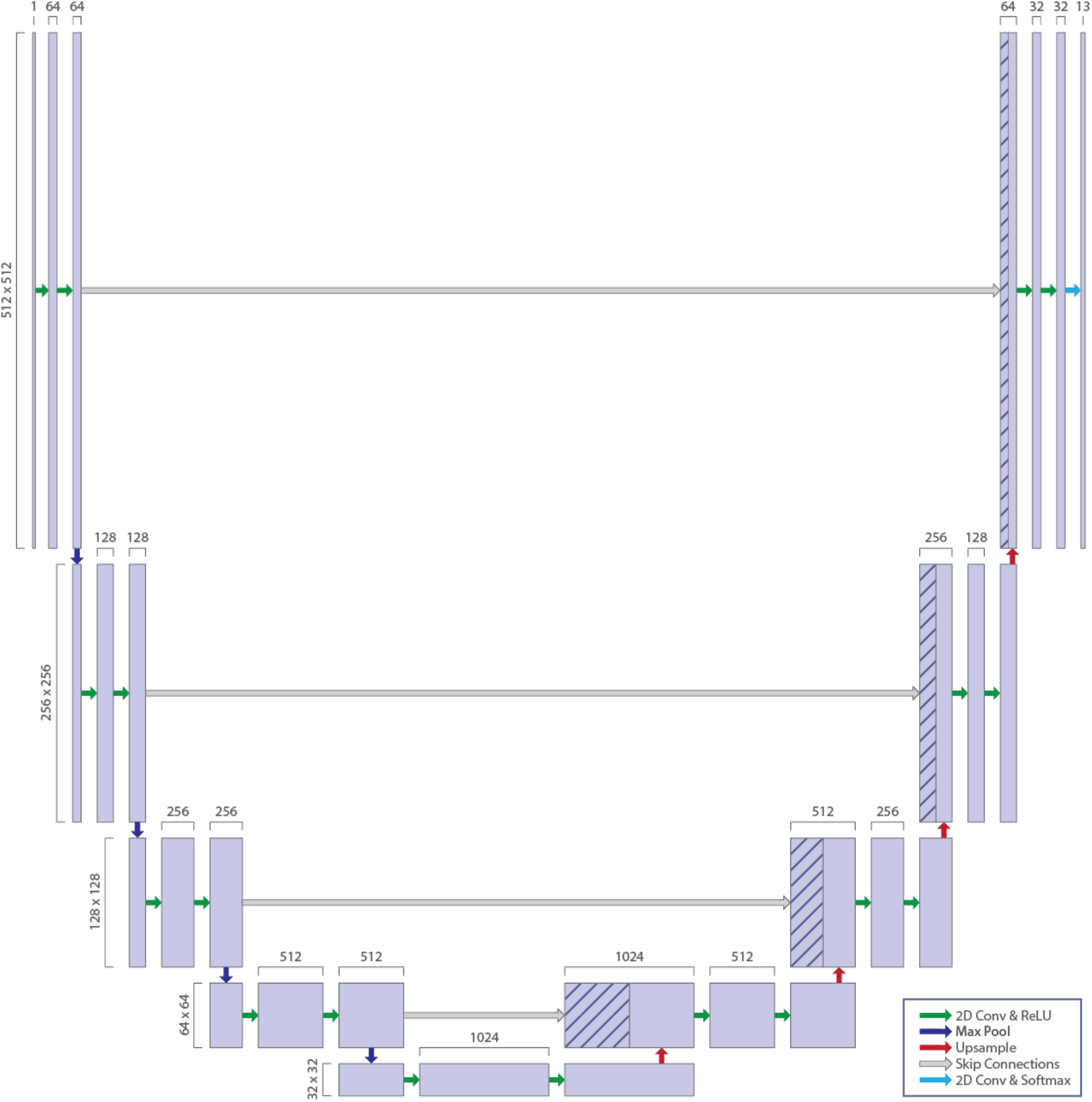
U-Net neural network architecture for semantic segmentation of CT images.

**Figure S2:**
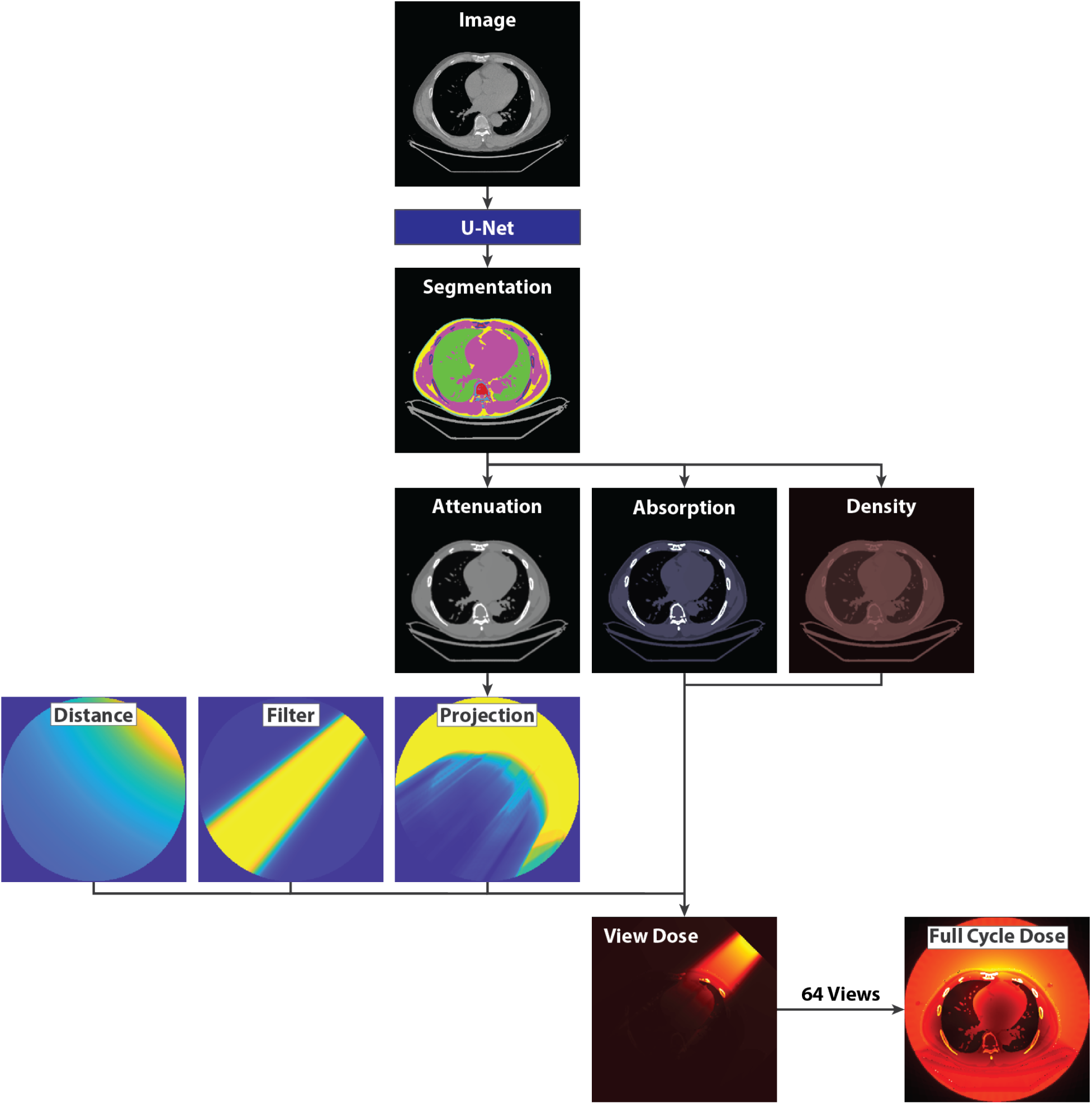
Dose simulation pipeline. The CT images are segmented using a U-Net. Material properties (linear attenuation, linear absorption, and mass density) are assigned according to tissue types. Linear attenuation values are integrated to form the projection which is combined with a filter attenuation profile and inverse-square distance from the source to compute the dose at a given view angle. These view doses are integrated at 64 angles to compute the full cycle dose map.

**Figure S3:**
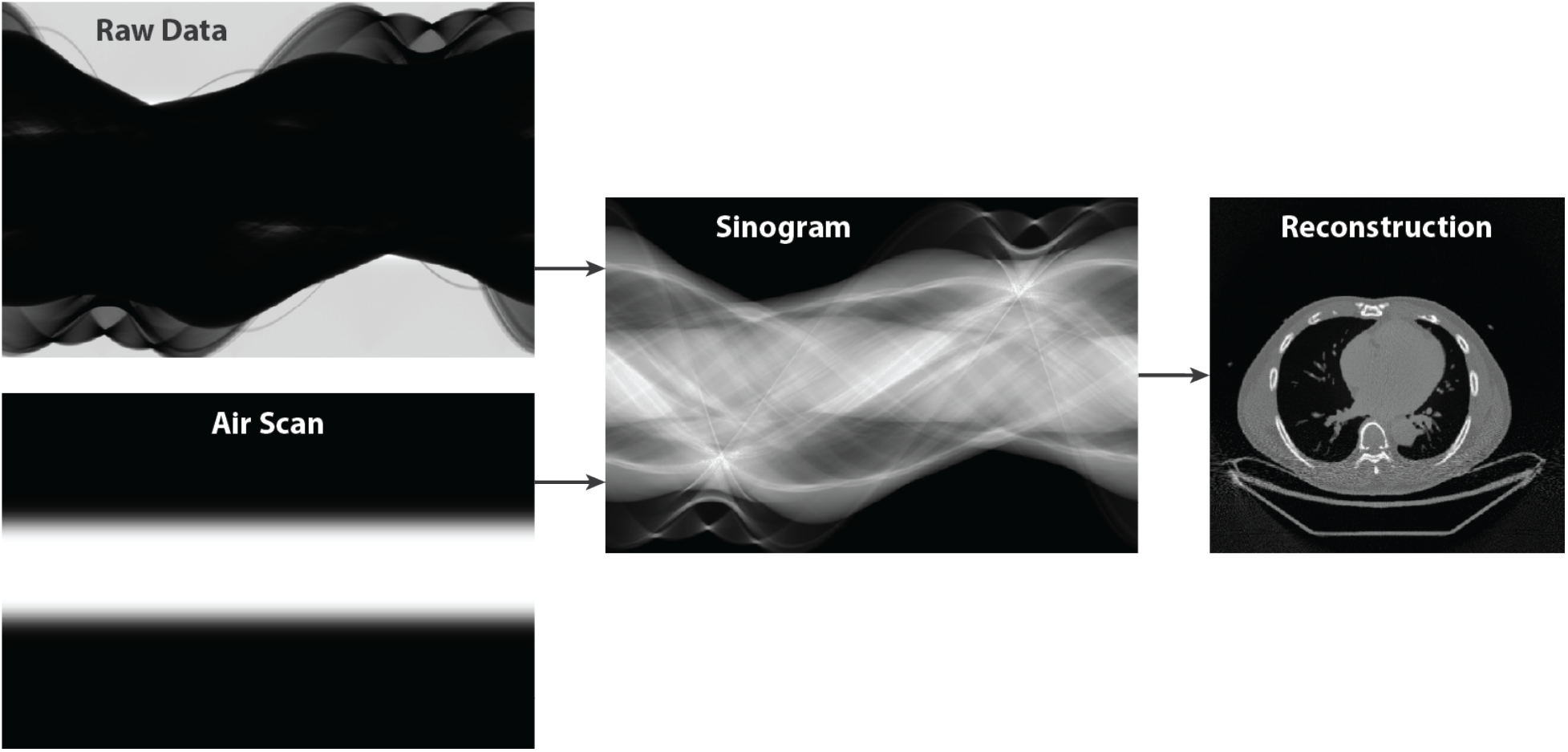
Image simulation pipeline. The central slice of the CT imaging process is simulated. First, a raw count and air scan is computed, then the sinogram is created. The sinogram is finally reconstructed using fanbeam filtered backprojection.

**Figure S4:**
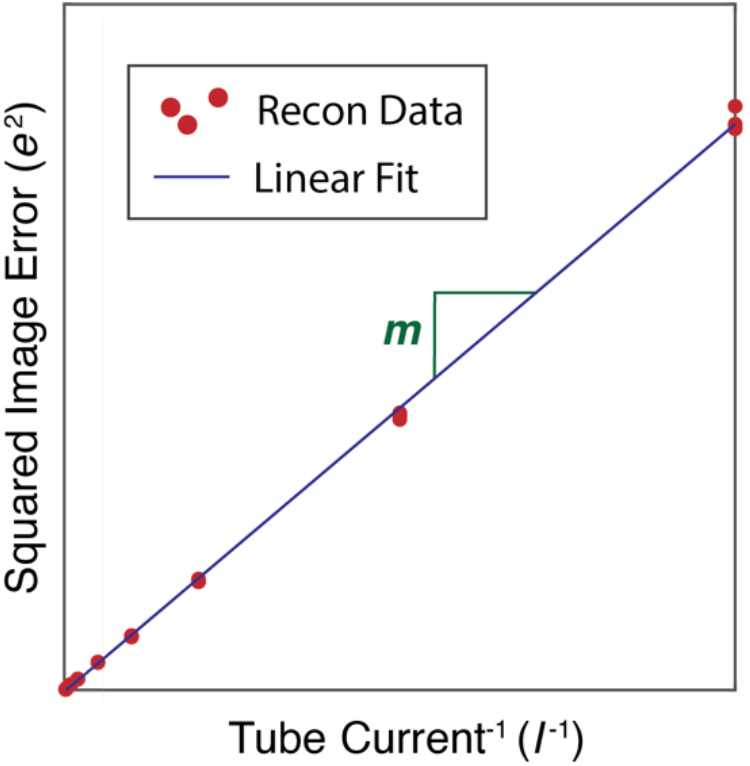
Image noise quantification. Image noise is quantified by the slope of a linear fit of observations of the relationship between tube current and reconstructed image error. The lower this value is, the better the image quality is for a given fluence level.

